# Gastrointestinal symptoms as Covid-19 onset in hospitalized Italian patients

**DOI:** 10.1101/2020.04.20.20064873

**Authors:** Elisabetta Buscarini, Guido Manfredi, Gianfranco Brambilla, Fernanda Menozzi, Claudio Londoni, Saverio Alicante, Elena Iiritano, Samanta Romeo, Marianna Pedaci, Gianpaolo Benelli, Ciro Canetta, Giuseppe Lapiana, Guido Merli, Alessandro Scartabellati, Giovanni Viganò, Roberto Sfogliarini, Giovanni Melilli, Roberto Assandri, Daniele Cazzato, Davide Sebastiano Rossi, Susanna Usai, Irene Tramacere, Germano Pellegata, Giuseppe Lauria

## Abstract

**Objective:** To assess the prevalence of gastrointestinal symptoms and their correlation with need of non-invasive ventilatory support, intensive care unit admission and death in hospitalized SARS-CoV-2 patients.

**Design:** Since February 21^th^ 2020, all individuals referred to our emergency department for suspected SARS-CoV-2 underwent a standardized assessment of body temperature and pulse oximetry, hematological screening, chest X-ray and/or computed tomography (CT), and SARS-CoV-2 assay on nasopharyngeal swab. Medical history and GI symptoms including nausea, vomit, diarrhea, and abdominal pain were recorded. were recorded.

**Results:** GI symptoms were the main presentation in 42 (10.2%) of 411 patients, with a mean onset 4.9 +/-… days before admission. In 5 (1.2%) patients GI symptoms have not been associated with respiratory symptoms or fever. We found an inverse trend for ICU admission and death as compared with patients without GI symptoms.

**Conclusions:** GI symptoms can be an early and not negligible feature of Covid-19, and might be correlated with a more benign disease course.

## Introduction

On January 7th, 2020, a novel coronavirus was isolated and named as severe acute respiratory syndrome coronavirus 2 (SARS-CoV-2) by the International Committee on Taxonomy of Viruses (ICTV) in the wake of an outbreak of pneumonia of unknown cause in Wuhan city, China.^1,2^ The World Health Organization (WHO) declared the 2019-nCoV outbreak a Public Health Emergency of International Concern on January 30^th^ 2020. This pneumonia was called coronavirus disease 2019 (Covid-19) on February 11th, 2010.The Covid-19 pandemic was declared on March 11^th^.

The better delineation and understanding of Covid-19 clinical features is of paramount importance to maximize diagnostic accuracy and timeliness, and to correlate clinical findings with outcome. It is well established that most patients with Covid-19 have fever along with respiratory signs and symptoms, such as cough and dyspnea.^3-5^ SARS-CoV-2 uses the ACE2 protein as a receptor, which is expressed not only in the respiratory epithelium, but also in the gastro-enteric mucosa.

A number of patients infected with SARS-CoV-2 is reported to present gastrointestinal (GI) symptoms including diarrhea, nausea, vomiting and/or abdominal pain or discomfort at onset or even before respiratory symptoms^6,7 8^

We investigated all consecutive individuals suspected to harbor Covid-19 and admitted at the General Hospital of Crema between February 21^st^ and March 13^rd^, 2020. This study is aimed at assessing prevalence and features of GI symptoms in Covid-19 patients and their correlation with medical history, disease course and outcome.

## METHODS

Since February 21^th^, all suspected individual admitted to the hospital underwent a standardized procedure including body temperature and pulse oximetry (SO_2_) recording, hematological screening, chest X-ray and/or computed tomography (CT) scan, and nasopharyngeal swab. Swabs were stored at +4°C and immediately shipped to one of the laboratory of virology accredited by the Lombardy Region for diagnostic SARS-COV-2 real-time polymerase chain reaction (RT-PCR) assay. Based on clinical, laboratory, and radiological findings, patients were discharged to home in quarantine or hospitalized.

Demographic data, date of onset and type of symptoms, including GI symptoms (as either nausea, or vomit or diarrhea or abdominal pain) were recorded. Comorbidities (hypertension, cardiovascular disorders, diabetes, pulmonary diseases, active and previous malignancies, any other disease), current pharmacological treatments and number of drugs were also recorded. All available clinical data during the hospitalization, including hematological and radiological exams, treatments, need of respiratory support with continuous positive airway pressure (CPAP) or non-invasive ventilation (NIV), ICU admission, and death were recorded. Follow-up data until March 19^th^ were recorded. Interstitial pneumonia was diagnosed based on acute reticular pattern chest X-ray and/or single or multiple ground-glass and/or consolidative lungs opacities at computed tomography (CT). Pleural and pericardial effusion, and lymphadenopathy at CT scan were recorded. Primary outcomes were need of CPAP or NIV, intensive care unit (ICU) admission, and death.

The hospital is equipped with a computerized recording system that generates a unique code for any visit and exam. All patients were anonymized and locked to the unique code assigned at the admission, and all data were included in an electronic database.

Steering Committee for the COVID-19 studies at ASST Maggiore Hospital Crema approved the study, which was notified to the Ethical Committee of ATS ValPadana.

### Statistical analysis

Descriptive statistics were provided in terms of absolute number and percentage for categorical data, and mean with standard deviation (SD) and value range for continuous data. Associations between GI symptoms and medical history, disease course and outcome were assessed through the use of chi-square test.

## RESULTS

### Clinical features and outcomes

We collected relevant data from 411 consecutive Covid-19 patients. Among them, 42 (10.2%,15 females and 27 males, mean age 68.2±14.2) reported GI symptoms including nausea (18, 4.3%), vomit (16, 3.8%),diarrhea (15, 3.6%) or abdominal pain (5, 1.2%); the 369 Covid-19 patients not reporting GI symptoms were 118 females, 251 males, mean age 67.6±15.0.

GI symptoms had a mean onset of 4.9 ±4.4 days (range 1-20) before admission. Patients with GI symptoms reported less frequently cough (p=0.004) as compared to those presenting with respiratory symptoms, whereas the frequency of fever was similar (p= 0.7). In 5 of 411 patients (1.2%) GI symptoms were neither associated with fever nor cough.

GI symptoms did not show any significant correlation with syncope (p=0.3), use of ACE inhibitors (p=0.1), presence of comorbidities (p=0.3), or use of multiple drugs (p=0.7).

Of the 42 patients presenting with GI symptoms, 9 (21.4%) required CPAP/NIV, 1 (2.3%) was admitted to ICU, and 4 (9.5%) died.

Table 1 shows correlation of GI symptoms with outcomes as either CPAP/NIV or ICU admission or death.

**Table 1.**
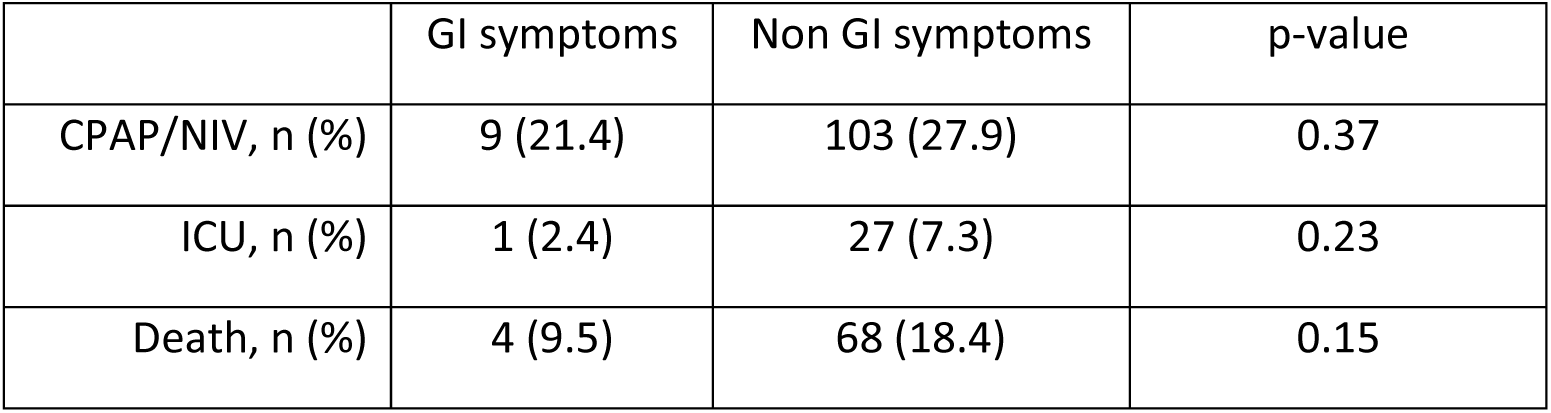

## DISCUSSION

Early observations in smaller cohorts reported that Covid-19 could present with GI symptoms, including diarrhoea, anorexia/nausea, abdominal discomfort and vomiting, in 3% of patients.^5 3^ Our finding of 10% patients suggests that the prevalence of GI symptoms at onset is not negligible and confirms the rate described in a Chinese cohort outside Wuhan.^9^

GI symptoms presented on average 4.9 days before admission. However, the range of GI symptoms onset is very wide with up to 20 days before admission, confirming that GI complaints can be the first herald of Covid-19.

Our data confirm the importance of including GI symptoms among the spectrum of Covid-19 features, to allow early diagnosis and appropriate treatments even in patients without respiratory symptoms. This could be of particular importance considering the rapid human-to-human transmission among close contacts, which could be related to GI rather than respiratory symptoms in some infected patients.^3^ Viral RNA is detectable in the stool of patients with suspicion of Covid-19. Gastrointestinal viral infection and potential fecal-oral transmission could persist even after viral clearance from the respiratory tract.^10,11^ As a consequence, prevention of oral-fecal transmission should be considered to control the spread of the virus.

In our cohort, GI symptoms did not correlate with fever, syncope, use of ACE inhibitors, comorbidities or use of multiple drugs. Conversely, we found a strong correlation with the absence of cough, suggesting that GI involvement might have an inverse correlation with lung involvement.

A possibly more benign disease course in patients with GI symptoms was suggested by the trend of ICU admissions and deaths, which was lower compared to patients without GI symptoms at onset, even though the difference was non significant possibly because of the relative small number of patients.

In conclusion, our study provided a reliable sizing of GI symptoms as early features of Covid-19 infection, strengthening the need to increase the awareness on potential orofecal transmission and widening the spectrum epidemiological constraints.

## Data Availability

The data that support the findings of this study are available from the corresponding author.

## Contributors

Study concept and design: EB, GL, GM. Data collection and statistical analysis: GM, RA, MP, IT. Analysis and interpretation of data: all authors. Drafting the manuscript: EB, GL, GM, IT. Critical revision of the manuscript for important intellectual content: all authors.

## Patient consent for publication

Not required

## Provenance and peer review

Not commissioned; externally peer reviewed

## Data availability statement

The data that support the findings of this study are available from the corresponding author.

